# Diagnostic yield of genetic screening in a diverse, community-ascertained cohort

**DOI:** 10.1101/2022.12.20.22283637

**Authors:** Nandana D. Rao, Jailanie Kaganovsky, Emily Malouf, Sandy Coe, Jenna Huey, Darwin Tsinajinne, Sajida Hassan, Kristine King, Stephanie M. Fullerton, Annie T. Chen, Brian H. Shirts

## Abstract

**Background:** Population screening for genetic risk of adult-onset preventable conditions has been proposed as an attractive public health intervention. Screening unselected individuals can identify many individuals who will not be identified through current genetic testing guidelines.

**Methods:** We sought to evaluate enrollment in and diagnostic yield of population genetic screening in a resource-limited setting among a diverse population. We developed a low-cost, short-read next-generation sequencing panel of 25 genes that had 98.4% sensitivity and 99.98% specificity compared to diagnostic panels. We used email invitations to recruit a diverse cohort of patients in the University of Washington Medical Center system unselected for personal or family history of hereditary disease. Participants were sent a saliva collection kit in the mail with instructions on kit use and return. Results were returned using a secure online portal. Enrollment and diagnostic yield were assessed overall, and across race and ethnicity groups.

**Results:** Overall, 40,857 people were invited and 2,899 (7.1%) enrolled. Enrollment varied across race and ethnicity groups, with the lowest enrollment among African American individuals (3.3%) and the highest among Multiracial or Other Race individuals (13.0%). Of 2,864 enrollees who received screening results, 106 actionable variants were identified in 103 individuals (3.6%). Of those who screened positive, 30.1% already knew about their results from prior genetic testing. The diagnostic yield was 74 new, actionable genetic findings (2.6%). The addition of more recently identified cancer risk genes increased the diagnostic yield of screening.

**Conclusions:** Population screening can identify additional individuals that could benefit from prevention, but challenges in recruitment and sample collection will reduce actual enrollment and yield. These should not be overlooked in intervention planning or in cost and benefit analysis.

## Background

Between 1% and 2% of the population has a genetic variant conferring a high lifetime risk of cancer or cardiovascular disease that can be mitigated by early application of screening, lifestyle changes, or medical intervention.^1–4^ Despite the availability of clinical testing and useful prevention methods, over 85% of people who are detected to have hereditary cancer risk have already developed cancer.^5–7^ Similarly, the majority of people with hereditary hypercholesterolemia find out about their disease predisposition later in adulthood.^8^ Identifying at-risk individuals early on is a challenge. Many at-risk individuals do not have family history of disease, most physicians and health systems do not have a process to routinely screen family history for genetic testing criteria, and limited access to genetic counseling may prevent individuals with family history from seeking testing.^1,9,10^ Cascade screening starting with individuals who have already been diagnosed could theoretically identify all at-risk individuals,^11^ but there are many barriers to effective family outreach.^12,13^

Expanding genetic screening to unselected members of the population has been proposed as a strategy to identify individuals who would benefit from timely medical intervention.^14,15^ The potential of population screening has been illustrated by studies that have implemented population screening using biobank samples in cohorts recruited for genomic research where as many as 75% of positive results were in individuals who would not have been identified through current genetic testing guidelines.^1–4^ However, the effectiveness of population screening among those who are not already enrolled in genetic research has not been thoroughly studied. One Israeli study of *BRCA1* and *BRCA2* screening among Ashkenazi Jewish individuals found that healthcare recruiters achieved between 55% and 92% enrollment depending on the study site and recruitment strategy.^16^

The diagnostic yield of invitation-based screening for genetic risk of preventable disease using a panel testing approach has also not been reported. Here we define diagnostic yield of population genetic screening as the number of individuals with newly-identified actionable findings. Many factors can influence diagnostic yield including the size, sensitivity, and specificity of the screening panel and the pretest characteristics of the target population specified in the design of the intervention. Including more genes in a screening panel is expected to increase yield. The Israeli and MyCode biobank screening studies included three variants and the entire *BRCA1* and *BRCA2* genes, respectively. The more recent Healthy Nevada Project and several ongoing screening studies have opted to use broader genetic screening panels.^1,3,4,17^ Recruitment strategy can also influence yield. Provider-referral to genetic screening has been associated with higher participation than self-referral; however, self-referral may have higher diagnostic yield due to higher likelihood of personal and family history of the conditioned screened among self-referred.^4,16^ Redundant testing, which may or may not be avoidable, reduces meaningful yield; MyCode and Nevada-based screening trials reported that positive results were already available to 18% and 12% of screened participants, respectively^1,2^

We sought to conduct population genetic screening for common, preventable inherited disease among a group of adult University of Washington Medicine (UWM) patients. The mandate of the study funders was to develop an effective, low-cost screening panel of 20 to 30 genes including hereditary cancer and hyperlipidemia; then recruit and screen over 2,500 individuals enriched for social and ethnic minorities in two years starting in August 2019. Time and financial constraints precluded extensive community outreach or multi-arm trials. Thus, a feature of this study is that it may reflect the exigencies of public health programs more accurately than prior studies of population screening.

Understanding the feasibility and effectiveness of population genetic screening in a diverse population is dependent on both the enrollment and yield among those who may be less likely to receive genetic testing with the current barriers to healthcare.^18^ Our goals were to identify the baseline enrollment and yield of genetic screening among a diverse group in a limited resource setting, which contrasts with previously published population genetic screening studies performed in the context of prior biobank recruitment and consent. Study enrollment and drop out were assessed overall and across race and ethnicity. Results may guide future public health population genetic screening efforts and inform appropriate cost effectiveness evaluations.

## Methods

### Participants

Participants for genetic screening were identified through a UWM medical record search. Potential participants were limited to adults 25 to 60 years old who had visited UWM hospitals or clinics at least two times in the last 5 years (2015-2020) to increase the chances that participants would have access to follow up care. UWM includes a tertiary/quaternary care academic hospital, two community hospitals, a safety-net hospital providing much of the uncompensated care in the region, and a network of primary care clinics throughout Western Washington.

Patients who self-identified as racial and ethnic minorities in the electronic health record (EHR) were preferentially selected for recruitment to assess the feasibility of population genetic screening among a diverse population. Exact enrollment goals for self-identified racial and ethnic minorities were defined by our partners to meet the needs of a separate project and were 10% African American, 46% Asian American, 10% Hispanic, 6% Native American or Pacific Islander, 20% White and 8% identifying as Other or Multiracial. In order to ensure input from sexual and gender minorities 1,000 invitations were sent to individuals self-identifying as LGBTQ+ according to EHR records.

Individuals were excluded if their coded EHR information indicated previous orders for hereditary cancer panel testing ordered through the University of Washington. However, no detailed medical record review was conducted to exclude individuals who may have received genetic testing prior to entry into the University of Washington health system or testing not recorded formally in the EHR. No other specific health profiles were selected for inclusion or exclusion. Study invitations were sent via email and enrollment forms were only available online and in English. The study was approved by the University of Washington IRB (00009032).

### Protocol

#### Recruitment Email Invitations

Study recruitment, data collection, and enrollment steps are depicted in Figure S1 (Additional File 1). Study invitations and subsequent surveys were sent from June 2020 to July 2021 using REDCap. Email invitations contained an introductory description to the study and a link to additional study information. The link led to study information and frequently asked questions (FAQ) (see Additional File 1), which included details about genetic screening and study consent. Individuals who did not enroll in screening were sent up to two reminder emails about study participation.

#### Baseline Data Collection (T0), DNA Kit Request, and Enrollment

At the end of the FAQ, interested persons were asked if they were willing to continue to answer an initial survey (T0) containing 24 close-ended questions related to personal and family medical history, factors influencing decision-making about genetic screening, and intent to share results. After the survey, people were asked if they would like receive an at-home DNA sample collection kit. People who declined were asked if they would voluntarily share their reason(s) at several points of the online enrollment process.

Those who requested DNA screening were sent an Oragene OGR 500 saliva collection kit, written and emailed electronic study consent forms, and a stamped return envelope. If completed kits and consent forms were not returned within three weeks, up to three reminder emails were sent to encourage completion. In some instances, a phone call from the study coordinator replaced the final, reminder email.

Study enrollment was complete once both the DNA kit with a collected sample and signed consent forms were returned. Invitees include all people who were sent study invites while enrollees include those who provided DNA samples and signed consent forms.

#### Post DNA Kit Return Data Collection (T1)

Once DNA kits were received in the laboratory, the samples were sequenced (See Table S1 for list of genes sequenced and Supplemental Methods for assay information and sequencing details in Additional File 1). Enrollees were notified of kit receipt at the lab via email and asked to complete a second online survey (T1), which asked more detailed questions about personal and family medical history, demographics, intent to share results, and knowledge about genetics. The T1 survey also included questions about prior genetic testing, bone marrow transplants, and others in their family who had received genetic testing results. Up to two reminder emails were sent to encourage completion.

#### Results Return

Enrollees whose test indicated no increased risk for the screened conditions were notified that their results were available via email and were able to access their result letter for viewing and printing on a secure and private website using two identifiers: a code uniquely assigned to them and their birthdate. Importantly, the result letter stressed that their risk was not known to be increased compared to the general population. The letter further emphasized that enrollees should consult with their physician for personal prevention recommendations.

A study genetic counselor contacted enrollees whose test indicated increased risk to schedule a phone conversation. The enrollee’s result letter was posted on the secure website described previously at the time of genetic counseling. Genetic testing results were not automatically included in the EHR, and enrollees were encouraged to share their results with their medical provider and obtain confirmatory clinical testing. Both the consent form and result letter stressed that medical management and/or follow-up would not occur within the research study. Study genetic counselor and faculty were available for additional consultations with the enrollee or their provider, if requested.

#### Post Result Return Data Collection (T2)

Enrollees received an email request to complete an optional post-results survey (T2) after they were sent their genetic screening results. The T2 survey consisted of both close- and open-ended items and asked about plans to share screening results, future health plans, and feelings about results and genetic screening. Up to two reminder emails were sent to encourage completion.

### Data Analysis

We used descriptive statistics to evaluate enrollment, yield, and result return. We used logistic regression to examine the association between race and ethnicity and enrollment. Model A included age, gender, and race/ethnicity with “Asian” used as reference. Model B added a gender by race/ethnicity interaction term. We also conducted an exploratory analysis to assess the relationship of sexual orientation with study enrollment by adding sexual orientation and a sexual orientation by race and ethnicity interaction term to Model A. We used self-reported personal and family history, including history of prior genetic testing to determine if enrollees were likely to have qualified for genetic testing under National Comprehensive Cancer Network guidelines for testing cancer-risk genes or if they would have qualified for familial hypercholesterolemia testing under American College of Cardiology guidelines.^19,20^ Because personal and family history were self-reported and the study did not have access to complete medical records, it was only possible to evaluate a few of the guideline criteria. Comparisons of self-report of testing between of groups receiving positive and uninformative results were performed using Fishers exact test. Additional details about data coding and analysis are provided in a Supplemental Methods section of Additional File 1.

## Results

### Study Demographics

A total of 40,857 people were invited to enroll in the population genetic screening research study conducted at UWM, with 2,889 (7%) people enrolling. Demographics of invitees and enrollees are shown in Table 1. While 54% of invitees were female, 60% of enrollees were female. Within enrollees, 12% were African American, 57% Asian, 6.2% Native American, 0.8% Multiracial or Other Race, and 23% White. Detailed information about enrollees, including health history, education, and income are listed in Table S2 (Additional File 1). Among enrollees, 53% of people had a family history of a cancer diagnosis, 61% reported having a college or advanced degree, and 38% reported a household income greater than $100,000.

**Table 1:** Electronic health record demographics of study invitees and enrollees.

|  | Invitees | Enrollees |
| --- | --- | --- |
| **N** | 40857 | 2889 |
| **Age<sup>a</sup>** | 39.6 (10.2) | 40.3 (10.1) |
| **Gender<sup>b</sup>** |  |  |
| Female | 22207 (54.3) | 1720 (59.5) |
| Male | 18537 (45.4) | 1099 (38.1) |
| Other | 81 (0.2) | 50 (1.7) |
| Prefer not to answer | 32 (0.1) | 20 (0.7) |
| **Race<sup>b</sup>** |  |  |
| African American | 10639 (26) | 354 (12.3) |
| Asian | 22043 (54) | 1646 (57.0) |
| Multiracial/Other | 177 (0.4) | 23 (0.8) |
| Native American | 2543 (6.2) | 182 (6.2) |
| White | 5437 (13.3) | 671 (23.2) |
| Missing | 18 (0.1) | 13 (0.5) |
| **Ethnicity<sup>b</sup>** |  |  |
| Hispanic | 4099 (10) | 369 (12.8) |
| Non-Hispanic | 36739 (89.9) | 2507 (86.8) |
| Missing | 19 (0.1) | 13 (0.4) |

<sup>a</sup>Mean (SD); <sup>b</sup>N (%)

### Screening Enrollment and Drop Out

Email invitations for screening were undeliverable in 1,612 (3.8%) instances, 87.2% of invitees did not click on the link in the study invitation, and 13.4% did not request a DNA kit for sample collection after reading more about the screening study (Figure 1). Dropout was also seen after kits for sample collection were sent, with 35.8% not completing collection or signing consent forms. In total 2,864 individuals were sent screening results (7% of those invited and 99% of those that returned kits and consent). A study genetic counselor was able to speak with 102/103 (99%) individuals receiving positive screening results. For individuals receiving uninformative screening results, 2,399/2,761 (87%) accessed their result letter online.

**Figure 1:**
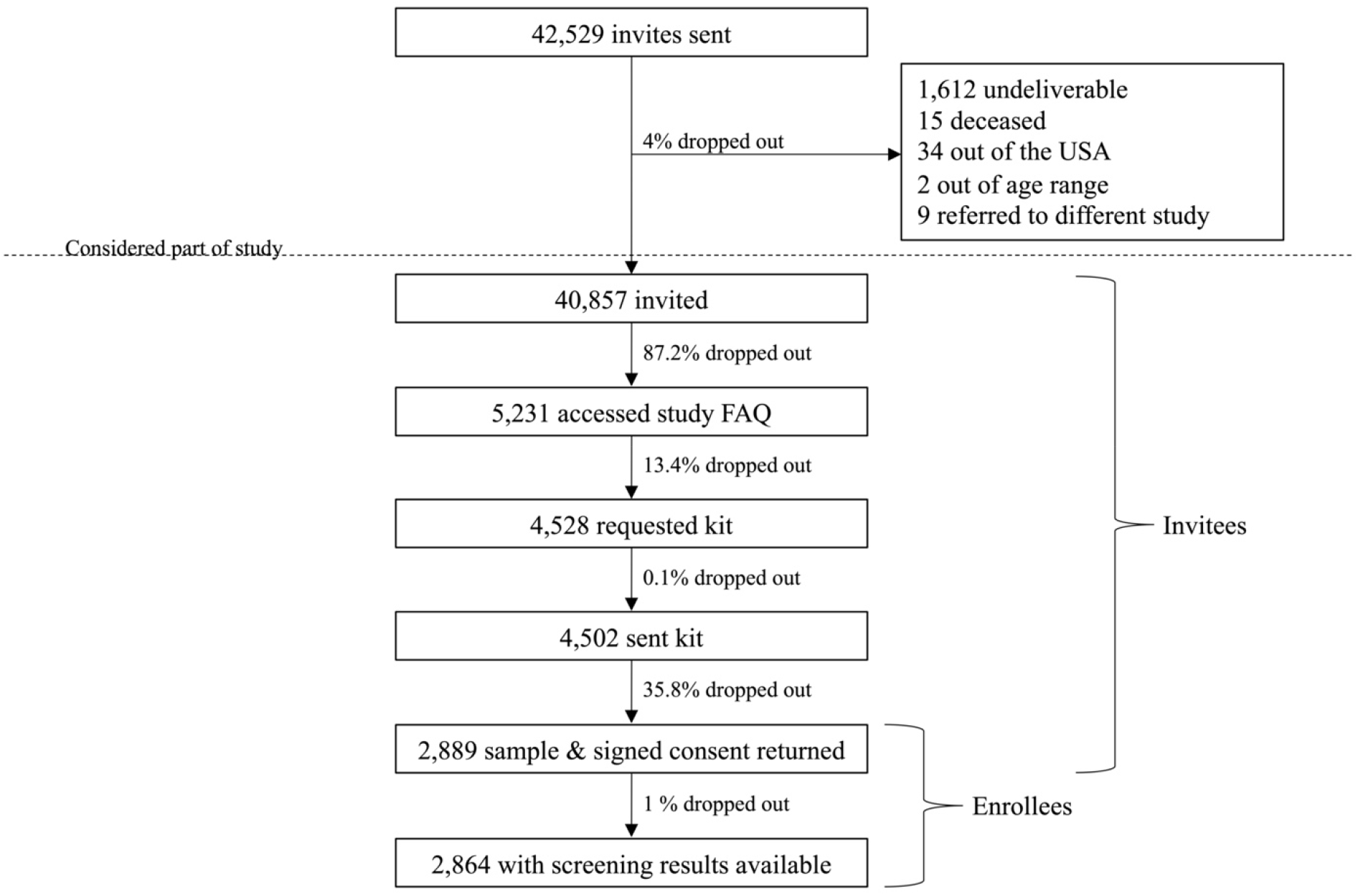
Study participation at different stages of population genetic screening. Flow diagram showing the enrollment and dropout of study subjects. “Dropout” at the last stage may have been due to technical failures or failure to return results to participants.

Trends in study involvement and dropout for all invitees and across race and ethnicity groups are shown in Figure 2 and are detailed in Table S3 (Additional File 1). African American individuals had the lowest rate of accepting the invitation to learn about the study compared to other groups, and a higher percentage of the small Multiracial or Other identifying individuals group continued to access the study FAQ compared to other groups. Between when DNA sample collection kits and consent forms were sent out and returned, the greatest dropout was seen for Multiracial or Other Race individuals and Native American individuals.

**Figure 2:**
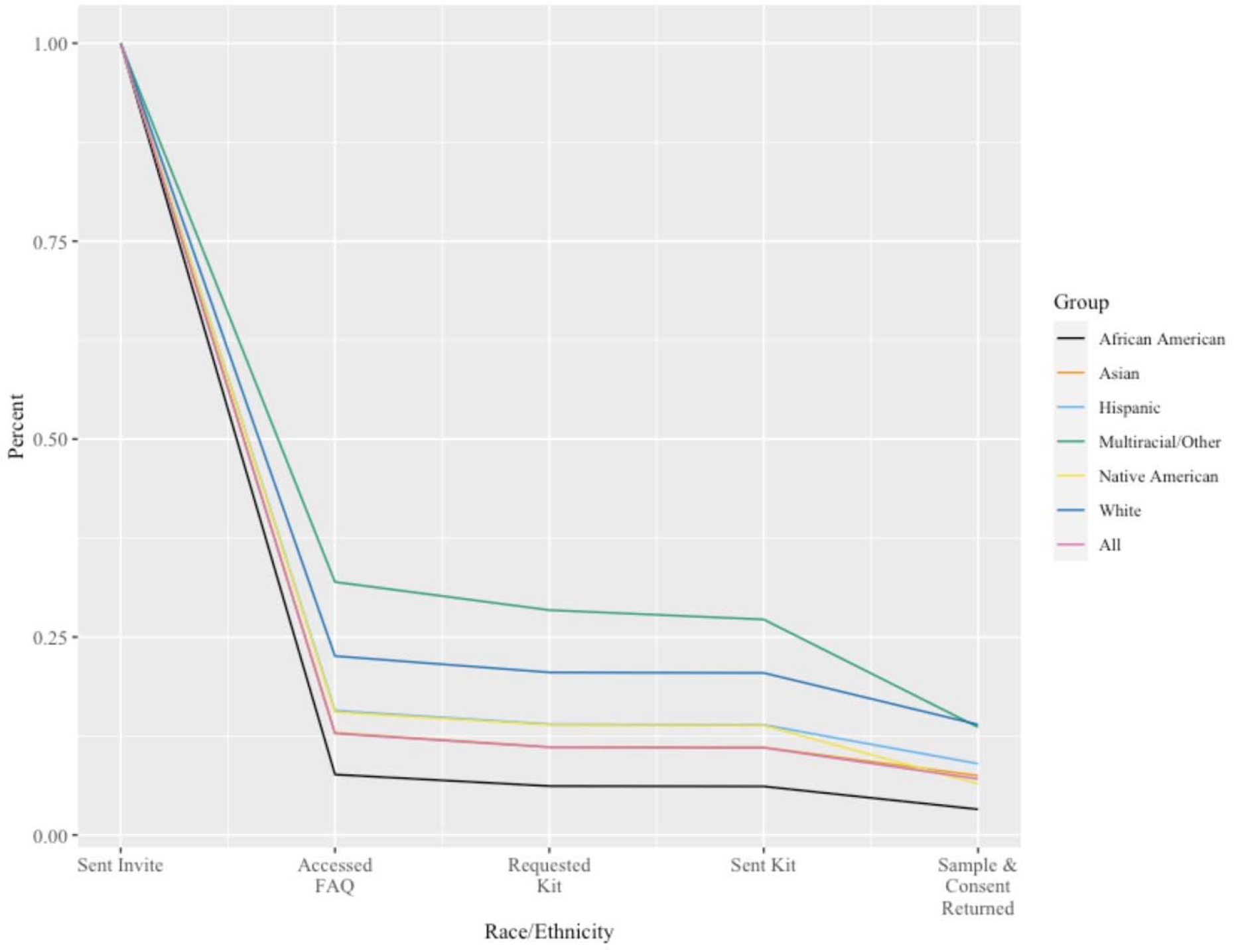
Study drop out by race and ethnicity. Lines trace the percentage of each group remaining in the study cohort at five stages of the study. The entire group is represented by the red line.

After adjusting for age and gender, results from logistic regression Model A (Table S4, Additional File 1) showed that African American individuals had lower odds of enrollment compared to Asian individuals (OR: 0.41, 95% CI: [0.36, 0.46]). White, Multiracial or Other Race, and Hispanic individuals had higher odds of enrollment compared to Asian individuals (OR: 1.81, 95% CI: [1.61, 2.04]; OR: 1.75, 95% CI: [1.1, 2.79]; OR: 1.21, 95% CI: [1.07, 1.36], respectively). Enrollment odds did not significantly differ between Native American and Asian individuals. An exploratory analysis adding sexual orientation and a sexual orientation by race and ethnicity interaction term to this model showed no independent association between sexual orientation and enrollment.

Results from Model B showed a significant interaction with male gender and African American race (p=0.004), such that odds of enrollment were lower for African American men compared to African American women (Table S5, Additional File 1). Similarly, a significant interaction was also seen with male gender and Hispanic ethnicity (p=0.008), such that odds of enrollment were lower for Hispanic men compared to Hispanic women.

### Screening Yield

Of the 2,864 enrollees who received results, 103 screened positive for at least one actionable variant (3.6%) (See Table S6 in Additional File 1 for complete list of variants identified). Positive results were reported for 17 unique genes (Figure 3). Three individuals had pathogenic variants in two genes (*ATM* and *BRIP1, BRCA2* and *CHEK2, BRCA2* and *LDLR*). The test panel identified 57 pathogenic or likely pathogenic variants in 9 genes associated with CDC tier-1 priority syndromes for genetic disease prevention^21^ (*BRCA1, BRCA2, MLH1, MSH2, MSH6, PMS2, EPCAM, APOB*, and *LDLR*) giving a yield of 2.0% for these genes. The panel identified 48 pathogenic or likely pathogenic variants in 10 other genes, most of which cause relatively lower lifetime risk of disease than tier-1 syndrome associated genes, giving a 1.6% carrier rate for this group of genes. No positive results were identified in the *BMPR1A, EPCAM, MUTYH, NTHL1, PTEN, SMAD4*, or *TP53* genes. The most frequently observed variant was reported in five individuals (*CHEK2* p.Ile157Thr), four variants were reported in three individuals (*CHEK2* 1100delC, *HOXB13* p.Gly84Glu, *LDLR* p.His583Tyr, and *RAD51C* c.394dup), and two variants reported in two individuals (*APC* p.Ile1307Lys and *APOB* p.Arg3527Gln).

**Figure 3:**
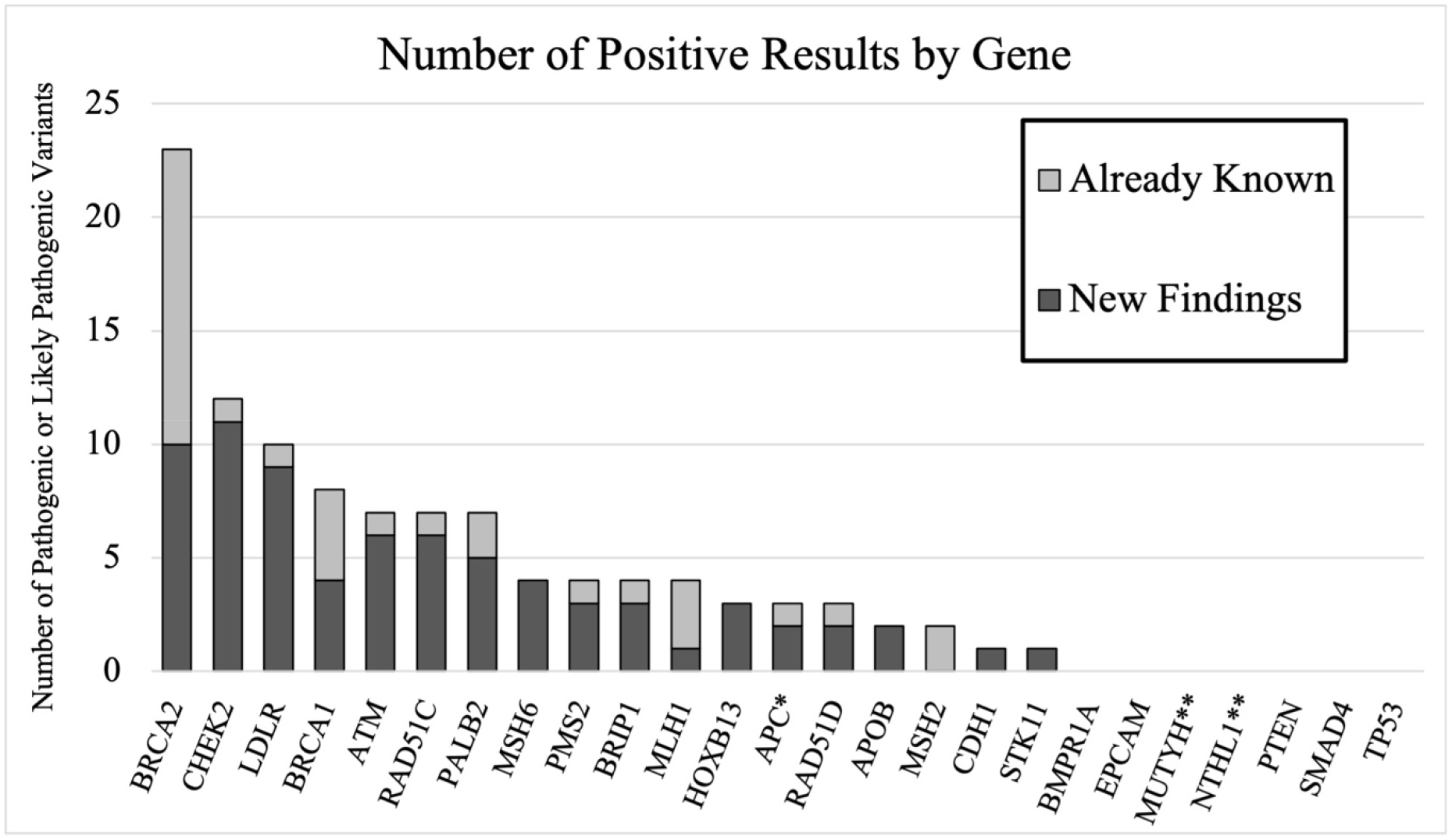
Screening results. Number of positive results by gene. Light grey bars show results that were already known by participants. Dark gray bars show new findings. *New findings were for APC p.Ile1307Lys, already know finding was for c.221-2A>G. **Study design was that only homozygous or compound heterozygous MUTYH and NTHL1 individuals were to be reported as positive.

Thirty-one of the 103 individuals (30.1%) who screened positive reported knowing about their results from prior genetic testing in the T1 survey (Table 2), although two of the three individuals who had two findings only knew about one of these (Table S6, Additional File 1). Of the 2,761 enrollees with uninformative results, 56 (2.0%) reported receiving prior sequencing for at least one of the genes on the screening panel, but none of the enrollees with uninformative results reported previous knowledge of having pathogenic or likely pathogenic variants in the genes tested. The frequency of reporting prior testing for genes in the panel was significantly different between those receiving positive and uninformative results (p < 0.0001). For the 9 genes associated with tier-1 syndromes, 42% (24/57) of pathogenic variants were already known reducing the true diagnostic yield from 2.0% to 1.2% for these genes. For other genes, 17% (8/48) of variants were already known, reducing diagnostic yield from 1.6% to 1.4% for this group of genes. Previously known variants were most likely to associated with tier-1 syndromes (p = 0.006). The overall diagnostic yield was 2.6%, with 74 new pathogenic or likely pathogenic variants identified in the 2,864 samples sequenced.

**Table 2:** Enrollee personal and family history of disease.

|  | **Aware of risk variant prior to screen** | **Known close relative\* with relevant variant** | **Personal diagnosis of disease (any age)** | **Close relative\* diagnosed with disease (any age)** | **Met any guidelines for testing** |
| --- | --- | --- | --- | --- | --- |
| **Enrollees with positive screening (n=103)** |  |  |  |  |  |
| HBOC | 23 | 16 | 12 | 33 | 16 |
| Lynch and polyposis | 7 | 4 | 5 | 7 | 5 |
| FH | 1 | 0 | NA | NA | 0 |
| **Enrollees with uninformative screening (n= 2761)** |  |  |  |  |  |
| HBOC | NA | 18 | 99 | 717 | 45 |
| Lynch and | NA | 6 | 30 | 345 | 59 |
| polyposis |  |  |  |  |  |
| FH | NA |  | NA | NA | 12 |

\* Parent, sibling, child

### Personal and family history of cancer and guidelines

Twenty individuals with positive results were aware of a pathogenic or likely pathogenic variant in a first-degree relative before enrolling in the study (19.4%) (Table 2). Of these individuals who knew about a close relative with their variant, screening genetic screening results were already known to 17 and new to three of the individuals (2.9% of all with positive results); one of these three received clinical testing after enrolling in the study and before receiving screening results. Only 24 people with uninformative screening results (0.9%) reported knowledge of a pathogenic variant in a first degree relative in study surveys. Of the 61 individuals who were likely to have met HBOC screening criteria, 16 had variants associated with increased risk of breast or ovarian cancer. Of the 64 individuals who were likely to have met Lynch syndrome or polyposis screening criteria, five had variants associated with increased risk colon cancer or polyposis. Of the 12 screened who were likely to have met personal and family history criteria for hypercholesterolemia screening criteria, none had variants associated with familial hypercholesterolemia. Overall 21 of 103 (20.5%) people with positive screening results and 116 of 2,761 (4.2%) people with uninformative screening results were likely to have met guidelines for diagnostic testing based on self-reported information from study surveys.

## Discussion

The methods used in this screening study for assay design; ascertaining, consenting, and enrolling participants; collecting samples; and disseminating genetic results may be representative of what a population genetic screening program might look like if implemented as a public health initiative by a resource-limited healthcare or government organization. Theoretically, every trial will be tailored to both the resources available and the needs of the community. Importantly, our strategy differed from prior trials of population genetic screening which were designed to determine yield of screening in an ideal situation using samples already collected, consented, and quality-controlled in a biorepository.^1–4^

Enrollment in our genetic screening trial was lower than that reported by previous population genetic screening studies. Among new participants of the Bio*Me* Biobank, 93% indicated that they wished to receive genetic results and more than 85% of people in the Geisinger MyCode biobank consented to participate in screening.^3,22^ It is possible participants in both biobank studies were already amenable to research participation and genetic testing leading to higher screening enrollment. Integration of screening into existing healthcare practices may also increase enrollment. The DNA10K, a population genetic screening program mediated by primary care providers, reported that 77% of patients had a genetic screening order placed by their provider.^23^ Results from our study may more closely reflect enrollment outcomes if screening were implemented among an unselected population as part of a stand-alone program and suggest that integrating programs with existing healthcare may be very important.

Implementing population screening is a challenging multi-step process. This study demonstrates that several issues still need to be addressed before participation is considered a viable option for many. Our results replicate historical patterns of differential use of genetic services^24-30^ and highlight the need to recognize geographic, socioeconomic, education, gender, sexual orientation, and racial and ethnic diversity. The Healthy Nevada Project and Alabama Genomic Health Initiative (AGHI) have engaged in a variety of outreach efforts using mass media, public events, multiple site enrollment and community partnerships and have seen success in recruiting individuals from different racial and ethnic backgrounds.^31,32^ On-going and long-term efforts to build trust through stakeholder engagement can also aid recruitment efforts and may be necessary for equitable enrollment.^33^ Dropout between when kits were sent and returned indicates that in-person collection or additional support may be necessary to mitigate difficulties with sample collection. Drop out during this period may also signal that screening is not a high priority for some individuals, despite initial interest. Our results indicate that screening programs which include recruitment and sample collection may be very different than screening programs that take advantage of already collected, high-quality DNA. Efforts to use recruiters, recruitment sites, or physicians to engage the community are likely to yield higher enrollment and also substantially increase the overall costs of screening.^16,17,34^

This study found that a large proportion of individuals with hereditary disease risk are missed under optimal guideline-based clinical testing, which is consistent with prior studies. Although we are not able to make direct comparisons, our data are consistent with findings that individuals with increased disease risk due to personal or family history may be more likely to self-refer to screening.^16^ In addition, we found that individuals who have already received prior, potentially redundant or overlapping, genetic testing may also be more likely to seek additional testing through screening interventions. It can be challenging to comprehensively determine who had and had not met prior genetic testing exclusion criteria with current EHRs,^35^ which may also limit selective screening in health systems. If not implemented thoughtfully and systematically, the reality may be that genetic screening will be like other screening tests that are repeated many times as technology improves and patients move between health systems.

The true diagnostic yield of 2.6% of this study was substantially lower than 3.6% carrier rate or total percentage of individuals who received positive results. We were surprised at the high rate of individuals who already knew about their genetic risk. This reduced the overall diagnostic yield substantially in comparison with the carrier rate. The diagnostic yield and carrier rate were both higher than those observed in the most similar prior study.^1^ The higher yield of our study was apparently due to several additional genes on the panel. If the comparison is limited to the 8 genes sequenced in both studies (*APOB, BRCA1, BRCA2, LDLR, MLH1, MSH2, MSH6*, and *PMS2*) the carrier rate of our study was higher (2.0%), but the final diagnostic yield of 1.2% was nearly identical. The similarity in yields between the two studies, despite demographic and recruitment differences, suggests that rare pathogenic variants may be represented at similar proportions in many populations.

There are many limitations to this study. Recruitment using only email is known to be less effective than other forms of recruiting. Our study only invited people for genetic screening who already had access to care in the UWM system. While this system includes a safety-net hospital, it may still underrepresent individuals without access to healthcare. Our enrichment for minorities may have produced results that are not representative of the general population. In addition, many of our analyses depend on EHR classifications of race and ethnicity; while these identifications are thought to be based on self-report, they may not be complete. We did not place screening results in the EHR, which may be a barrier to follow-up. As noted above, many limitations were judged to be acceptable in this pilot study as results would be more likely to mirror limitations of strategies that a resource-limited government or healthcare organization might undertake to implement population screening. The study is now following patients to monitor post-screening perceptions, barriers to follow-up care, and outcomes.

## Conclusions

Enrollment in population genetic screening in our diverse community-ascertained cohort for variants in 25 genes associated with preventable adult-onset disease was 7%, with a 2.6% diagnostic yield for individuals screened. While population screening has potential to identify additional individuals that could benefit from prevention, implementation challenges remain, particularly during recruitment and sample collection. These challenges to widespread adoption of population screening reduce actual enrollment and diagnostic yield and should not be overlooked in intervention planning or in cost and benefit analysis.

## Data Availability

Datasets generated and/or analyzed during the current study are not publicly available as they include some data from the University of Washington electronic health record and consent forms did not include broad data sharing. Deidentified datasets are available from the corresponding author on reasonable request.

