## Additional File 1 for "Diagnostic yield of genetic screening in a diverse, community-ascertained cohort"

### **Figure S1: Study protocol for population genetic screening**

Email invitation

Clicked email link to FAQ

Interested in continuing?

Yes

No

T0 Survey

Open text box to provide reason(s) for declining

Interested in receiving DNA kit?

Provide address info

Open text box to provide reason(s) for declining

Yes

No

DNA kits & consent forms sent

DNA kits & consent forms returned

T1 Survey

Result return

T2 Survey

Yes

No

### **Table S1: Full list of genes screened**

| **Gene** | **Associated Condition(s)** |
| --- | --- |
| APC | Colon polyps, colon cancer |
| APOB | High cholesterol, coronary artery disease |
| ATM | Breast cancer |
| BMPR1A | Colon polyps, colon cancer |
| BRCA1 | Breast, ovarian, and pancreatic cancer |
| BRCA2 | Breast, ovarian, and pancreatic cancer |
| BRIP1 | Breast and ovarian cancer |
| CDH1 | Breast and stomach cancer |
| CHEK2 | Breast and colon cancer |
| EPCAM | Colon, endometrial, and ovarian cancer |
| HOXB13 | Prostate cancer |
| LDLR | High cholesterol, coronary artery disease |
| MLH1 | Colon, endometrial, and ovarian cancer |
| MSH2 | Colon, endometrial, and ovarian cancer |
| MSH6 | Colon, endometrial, and ovarian cancer |
| MUTYH | Colon polyps, colon cancer |
| NTHL1 | Colon polyps, colon cancer |
| PALB2 | Breast and ovarian cancer |
| PMS2 | Colon, endometrial, and ovarian cancer |
| PTEN | Colon polyps, colon cancer |
| RAD51C | Ovarian cancer |
| RAD51D | Ovarian cancer |
| SMAD4 | Colon polyps, colon cancer |
| STK11 | Colon polyps, colon cancer |
| TP53 | Breast and many other cancers |

### **Table S2: Detailed demographics of enrollees**

| **Characteristic** | **N (%)** |
| --- | --- |
| **Adopted** |  |
| Yes | 115 (4.0) |
| No | 2762 (95.6) |
| Missing | 12 (0.4) |
| **Personal cancer diagnosis** |  |
| Yes | 321 (11.1) |
| No | 2560 (88.6) |
| Missing | 8 (0.3) |
| **Family cancer diagnosis**^a^ |  |
| Yes | 1535 (53.2) |
| No | 1024 (35.4) |
| Don’t know | 318 (11.0) |
| Missing | 12 (0.4) |
| **Personal heart attack**^b^ |  |
| Yes | 43 (1.5) |
| No | 2835 (98.1) |
| Missing | 11 (0.4) |
| **Family heart attack**^a^ |  |
| Yes | 1010 (34.9) |
| No | 1327 (46.0) |
| Don’t know | 545 (18.9) |
| Missing | 7 (0.2) |
| **Sex assigned at birth** |  |
| Male | 765 (26.4) |
| Female | 1295 (44.9) |
| Other | 0 (0) |
| Prefer not to answer | 3 (0.1) |
| Missing | 826 (28.6) |
| **Sexual orientation**^c^ |  |
| Asexual | 84 (2.9) |
| Bisexual | 147 (5.1) |
| Gay or lesbian | 172 (6.0) |
| Queer | 77 (2.7) |
| Straight | 1560 (54.0) |
| Something else | 20 (0.7) |
| Don’t know | 10 (0.4) |
| Prefer not to answer | 42 (1.5) |
| Missing | 841 (29.1) |
| **Education** |  |
| Less than high school | 6 (0.2) |
| High school/GED | 61 (2.1) |
| Some college | 299 (10.4) |
| College graduate | 865 (29.9) |
| Advanced degree | 887 (30.7) |
| Missing | 771 (26.7) |
| **Household Income** |  |
| <$50,000 | 304 (10.5) |
| >$50,000 but ≤100,000 | 539 (18.7) |
| >$100,000 | 1105 (38.2) |
| Prefer not to answer | 190 (6.6) |
| Missing | 751 (26.0) |
| **Past genetic testing** |  |
| Yes | 425 (14.7) |
| No | 1689 (58.5) |
| Missing | 775 (26.8) |
| **Family genetic testing showing increased risk**^d^ |  |
| Yes | 99 (3.4) |
| No | 1959 (67.8) |
| Missing | 831 (28.8) |

*^a^*Family was defined as a close biological relative (mother, father, son, daughter, aunt, uncle)

*^b^*At any point in time

*^c^*Multiple choices could be selected

*^d^*Degree of relatedness was not specified in question. This question was asked to all people administered the T1 survey and a not applicable option to indicate a lack of family genetic testing was not provided

**Table S3: Study participation at different stages of population genetic screening by race and ethnicity** [N (%)]

|  | **Overall** | **African American** | **Asian** | **Hispanic** | **Other** | **Native American** | **White** | **Missing** |
| --- | --- | --- | --- | --- | --- | --- | --- | --- |
| **Invited** | 40857 (100) | 10267 (100) | 21707 (100) | 4099 (100) | 169 (100) | 1533 (100) | 3063 (100) | 19  (100) |
| **Accessed FAQ** | 5231 (12.8) | 784  (7.6) | 2801 (12.9) | 644  (15.7) | 54 (32.0) | 238  (15.5) | 692 (22.6) | 18 (94.7) |
| **Requested kit** | 4528 (11.08) | 634  (6.18) | 2413 (11.12) | 573 (13.98) | 48 (28.4) | 213 (13.89) | 629 (20.54) | 18 (94.7) |
| **Sent kit** | 4502 (11.02) | 630  (6.14) | 2399 (11.05) | 570 (13.91) | 46 (27.2) | 212 (13.83) | 627 (20.47) | 18 (94.7) |
| **Sample & signed consent returned** | 2889 (7.1) | 333  (3.2) | 1625 (7.5) | 369  (9) | 23 (13.6) | 99  (6.5) | 427 (13.9) | 13 (68.4) |

### **Table S4: Association of race and ethnicity with enrollment**

| **Race/Ethnicity** | **Beta** | **SE** | **P-Value** | **OR (95% CI)** |
| --- | --- | --- | --- | --- |
| African American | -0.90 | 0.06 | <2e-16 | 0.41 (0.36, 0.46) |
| Hispanic | 0.19 | 0.06 | 0.002 | 1.21 (1.07, 1.36) |
| Multiracial/Other | 0.56 | 0.24 | 0.018 | 1.75 (1.1, 2.79) |
| Native American | -0.19 | 0.11 | 0.072 | 0.82 (0.67, 1.02) |
| White | 0.59 | 0.06 | <2e-16 | 1.81 (1.61, 2.04) |

*adjusted for age (continuous) and gender (male, female, other); Asian as reference

### **Table S5: Probability of screening enrollment for men and women aged 40 in race and ethnicity groups where a significant interaction was present with gender and race and ethnicity**

|  | **African American** | **Asian** | **Hispanic** |
| --- | --- | --- | --- |
| **Female** | 0.04 | 0.08 | 0.11 |
| **Male** | 0.03 | 0.07 | 0.07 |

Asian as reference

### **Table S6: Variants detected in the population genetic screening study**

| **Study ID** | **Gene and variant** | **Known to enrollee (Yes/No)** | **Known in first degree relative (Yes/No)** |
| --- | --- | --- | --- |
| 36534 | APC NM_000038.5:c.221-2A>G | Y | N |
| 14454 | APC NM_000038.5:c.3920T>A, p.Ile1307Lys | N | N |
| 44440 | APC NM_000038.5:c.3920T>A, p.Ile1307Lys | N | N |
| 32321 | APOB NM_000384.3:c.10580G>A, p.Arg3527Gln | N | N |
| 36032 | APOB NM_000384.3:c.10580G>A, p.Arg3527Gln | N | N |
| 5023 | ATM NM_000051.3:c.1564_1565del, p.Glu522Ilefs*43 | N | N |
| 2032 | ATM NM_000051.3:c.1939G>T, p.Glu647* | Y | N |
| 4694 | ATM NM_000051.3:c.3372C>G, p.Tyr1124* | N | N |
| 36465 | ATM NM_000051.3:c.5763-1050A>G | N | N |
| 23715 | ATM NM_000051.3:c.6347+1G>A | N | N |
| 18270 | ATM NM_000051.3:c.742C>T, p.Arg248* | N | N |
| 21727 | ATM NM_000051.3:c.8786+1G>A | N | N |
| 18588 | BRCA1 NM_007294.3 exon 9-12 del | Y | N |
| 22110 | BRCA1 NM_007294.3:c. 181T>G, p.Cys61Gly | Y | N |
| 44396 | BRCA1 NM_007294.3:c.188T>A, p.Lys63* | N | N |
| 38565 | BRCA1 NM_007294.3:c.191G>A, p.Cys64Tyr | N | N |
| 3023 | BRCA1 NM_007294.3:c.1960A>T, p.Lys654* | Y | N |
| 23736 | BRCA1 NM_007294.3:c.3756_3759del, p.Ser1253Argfs*10 | N | N |
| 15348 | BRCA1 NM_007294.3:c.5359_5363delinsAGTGA, p.Cys1787_Gly1788delinsSerAsp | Y | N |
| 26291 | BRCA1 NM_007294.3:c.5470_5477del, p.Ile1824Aspfs*3 | N | N |
| 1037 | BRCA2 NM_000059.3:c.1796_1800del, p.Ser599* | Y | N |
| 32904 | BRCA2 NM_000059.3:c.2059_2063del, p.Asp687* | N | N |
| 256 | BRCA2 NM_000059.3:c.2677del, p.Gln893Lysfs*2 | Y | N |
| 35852 | BRCA2 NM_000059.3:c.316+1G>A | Y | N |
| 2961 | BRCA2 NM_000059.3:c.3264dup, p.Gln1089Serfs*10 | Y | N |
| 747 | BRCA2 NM_000059.3:c.3554_3555del, p.Thr1185Serfs*2 | Y | N |
| 26195 | BRCA2 NM_000059.3:c.3860del, p.Asn1287Ilefs*6 | N | N |
| 20449 | BRCA2 NM_000059.3:c.4111C>T, p.Gln1371* | Y | N |
| 28913 | BRCA2 NM_000059.3:c.4449del, p.Asp1484Thrfs*2 | Y | N |
| 3696 | BRCA2 NM_000059.3:c.4588A>T, p.Lys1530* | N | N |
| 2087 | BRCA2 NM_000059.3:c.4631del, p.Asn1544Thrfs*24 | N | N |
| 35449 | BRCA2 NM_000059.3:c.5238dup, p.Asn1747* | N | N |
| 147 | BRCA2 NM_000059.3:c.5454del, p.Cys1820fs | Y | N |
| 37902 | BRCA2 NM_000059.3:c.5909C>A, p.Ser1970* | N | N |
| 1312 | BRCA2 NM_000059.3:c.6275_6276del, p.L2092Pfs*7 | Y | N |
| 18549 | BRCA2 NM_000059.3:c.6405_6409del, p.Asn2135Lysfs*3 | N | N |
| 36547 | BRCA2 NM_000059.3:c.658_659del, p.Val220Ilefs*4 | N | N |
| 2863 | BRCA2 NM_000059.3:c.6833_6837del, p.Ile2278fs | Y | N |
| 20886 | BRCA2 NM_000059.3:c.6952C>T, p.Arg2318* | Y | N |
| 43181 | BRCA2 NM_000059.3:c.7480C>T, p.Arg2494* | N | N |
| 30333 | BRCA2 NM_000059.3:c.7588C>T, p.Gln2530* | N | Y |
| 1392 | BRCA2 NM_000059.3:c.771_775del, p.N257Kfs*17 | Y | N |
| 20455 | BRCA2 NM_000059.3:c.8548_8551del, p.Glu2850Glnfs*12 | Y | Y |
| 2032 | BRIP1 NM_032043.2:c.2253_2254del, p.Lys752Argfs*12 | N | N |
| 48460 | BRIP1 NM_032043.2:c.2990_2993del, p.Thr997Argfs*61 | Y | N |
| 32796 | BRIP1 NM_032043.2:c.3201C>A, p.Cys1067* | N | N |
| 1983 | BRIP1 NM_032043.2:c.985C>T, p.Gln329* | N | N |
| 1164 | CDH1 NM_004360.3:c.1118dup, p.Ile374Aspfs*13, | N | N |
| 2892 | CHEK2 NM_007194.3:c.1100del, p.Thr367Metfs*15 | Y | N |
| 30137 | CHEK2 NM_007194.3:c.1100del, p.Thr367Metfs*15 | N | N |
| 32027 | CHEK2 NM_007194.3:c.1100del, p.Thr367Metfs*15 | N | N |
| 24950 | CHEK2 NM_007194.3:c.219_223del, p.Ile74* | N | N |
| 27967 | CHEK2 NM_007194.3:c.349A>G, p.Arg117Gly | N | N |
| 3740 | CHEK2 NM_007194.3:c.470T>C p.Ile157Thr | N | N |
| 2863 | CHEK2 NM_007194.3:c.470T>C, p.Ile157Thr | N | N |
| 6635 | CHEK2 NM_007194.3:c.470T>C, p.Ile157Thr | N | N |
| 21650 | CHEK2 NM_007194.3:c.470T>C, p.Ile157Thr | N | Y |
| 29897 | CHEK2 NM_007194.3:c.470T>C, p.Ile157Thr | N | N |
| 32415 | CHEK2 NM_007194.3:c.846+4del | N | N |
| 29954 | CHEK2 NM_007194.3:c.902del: p.Leu301Trpfs*3 | N | N |
| 13189 | HOXB13 NM_006361.6:c.251G>A, p.Gly84Glu | N | N |
| 16751 | HOXB13 NM_006361.6:c.251G>A, p.Gly84Glu | N | N |
| 40102 | HOXB13 NM_006361.6:c.251G>A, p.Gly84Glu | N | N |
| 36883 | LDLR NM_000527.4: exon 9-12 del | N | N |
| 16025 | LDLR NM_000527.4:c.1247G>A, p.Arg416Gln | N | N |
| 26472 | LDLR NM_000527.4:c.1424C>T, p.Ala475Val | N | N |
| 21330 | LDLR NM_000527.4:c.1426C>T, p.Pro476Ser | N | N |
| 34221 | LDLR NM_000527.4:c.1586+5G>A | N | N |
| 11952 | LDLR NM_000527.4:c.1747C>T, p.His583Tyr | Y | N |
| 35449 | LDLR NM_000527.4:c.1747C>T, p.His583Tyr | N | N |
| 41409 | LDLR NM_000527.4:c.1747C>T, p.His583Tyr | N | N |
| 45447 | LDLR NM_000527.4:c.1783C>T, p.Arg595Trp | N | N |
| 16068 | LDLR NM_000527.4:c.249del, p.Pro84Leufs*122 | N | N |
| 35110 | LDLR NM_000527.4:c.268G>A, p.Asp90Asn | N | N |
| 5861 | MLH1 NM_000249.3 exon 16-19 del | Y | N |
| 2063 | MLH1 NM_000249.3:c.1409+1G>A | Y | N |
| 510 | MLH1 NM_000249.3:c.245C>T, p.Thr82Ile | Y | N |
| 27469 | MLH1 NM_000249.3:c.793C>T, p.Arg265Cys | N | N |
| 13694 | MSH2 NM_000251.2:c.1835C>G, p.S612* | Y | N |
| 176 | MSH2 NM_000251.2:c.942+3A>T | Y | N |
| 5968 | MSH6 NM_000179.2:c.2759del, p.Lys920Argfs*25 | N | N |
| 34682 | MSH6 NM_000179.2:c.3119_3120del, p.Phe1040* | N | N |
| 42458 | MSH6 NM_000179.2:c.3259_3260insA, p.Pro1087Hisfs*6 | N | N |
| 3790 | MSH6 NM_000179.2:c.3261dup, p.Phe1088Leufs*5, | N | N |
| 1646 | PALB2 NM_024675.3:c.2167_2168del, p.Met723Valfs*21 | Y | N |
| 17611 | PALB2 NM_024675.3:c.2167_2168del, p.Met723Valfs*21 | Y | N |
| 2679 | PALB2 NM_024675.3:c.226del, p. Ile76Tyrfs*101 | N | N |
| 32148 | PALB2 NM_024675.3:c.3183_3184del, p.His1061Glnfs*5 | N | N |
| 19496 | PALB2 NM_024675.3:c.3244_3245del, p.Ser1082* | N | N |
| 19818 | PALB2 NM_024675.3:c.599del, p.Leu200* | N | N |
| 26231 | PALB2 NM_024675.3:c.751C>T, p.Gln251* | N | N |
| 16116 | PMS2 NM_000535.6:c.2444C>T, p.Ser815Leu | N | N |
| 960 | PMS2 NM_000535.6:c.2444C>T, p.Ser815Leu | Y | N |
| 37647 | PMS2 NM_000535.6:c.538-2A>G | N | N |
| 28464 | PMS2 NM_000535.6:c.614A>C, p.Gln205Pro | N | N |
| 1849 | RAD51C NM_058216.3:c.394dup, p.Thr132Asnfs*23 | N | Y |
| 30125 | RAD51C NM_058216.3:c.394dup, p.Thr132Asnfs*23 | Y | N |
| 40187 | RAD51C NM_058216.3:c.394dup, p.Thr132Asnfs*23 | N | N |
| 782 | RAD51C NM_058216.3:c.404+2T>C | N | N |
| 16669 | RAD51C NM_058216.3:c.577C>T, p.Arg193* | N | N |
| 19704 | RAD51C NM_058216.3:c.905-2A>C | N | N |
| 36277 | RAD51C NM_058216.3:c.97C>T, p.Gln33* | N | N |
| 23550 | RAD51D NM_002878.3:c.131_144+24del | N | N |
| 27492 | RAD51D NM_002878.3:c.270_271dup, p.Lys91leIfs*13 | N | N |
| 1380 | RAD51D NM_002878.3:c.694C>T, p.Arg232* | Y | N |
| 43871 | STK11 NM_000455.4:c.862+3G>T | N | N |

**Supplemental Methods:**

#### 1. **Screening assay design and validation**

The screening panel described to invitees consisted of genes associated with diseases or disorders of cancer and high lipids, for which clinical procedures or therapies are available for disease prevention, mitigation, and treatment. The testing panel was developed by UWM, Genetics and Solid Tumors Laboratory to detect pathogenic and likely pathogenic variants in the 25 covered genes including variants at intron/exon boundaries and single or multi-exon deletions or duplications. A list of the targeted genes and their associated conditions (Supplemental Table 1) was available on the study website. For two genes, *MUTYH* and *NTHL1*, it was decided to report only homozygous or compound heterozygous pathogenic and likely pathogenic variants.

DNA from Oragene saliva sample kits was isolated for sequencing at the UWM Genetics and Solid Tumors Laboratory or the UWM Northwest Clinical Genomics Laboratory using QIAsymphony automated extraction. DNA libraries were generated using IDT Lotus library prep kits followed by capture using a custom set of IDT biotinylated probes. Pools of between six and 188 enrollee samples with appropriate controls were sequenced on Illumina MiSeq or Next Seq sequencers. The bioinformatics pipeline for next-generation sequence analysis was a minimally-modified version of clinical UW ColoSeq assay pipeline and performance was expected to mirror performance of the clinical BROCA assay.^1,2^ Several unique customizations were incorporated into the bioinformatic pipeline, including processes to detect copy number variants in all exons of *PMS2*.^3^

Prior to the screening study, the panel was validated using 124 samples with 129 known variants of interest together with 9,404 variants from well characterized biobank samples. Across all variant types, the panel had 98.4% sensitivity for variants of interest, with lower sensitivity for indels greater than 12 bp and complex copy number variants, and 99.98% specificity for covered variants in biobank samples.

#### 2. **Data analysis**

*Screening enrollment and drop out*

Data on the process of genetic screening collected in a REDCap database included number of invites sent, number of DNA sample collection kits sent, enrollees, failed samples, and repeat testing. Demographic data, including age, gender, race, and ethnicity, were available for all study invitees through the UWM EHR. The T1 survey also asked about gender, race, and ethnicity. As T1 survey data were not reported for all invitees, EHR demographics were used for analyses to maximize available data. However, individuals who responded “Prefer not to answer” for the gender question in the T1 survey were classified as such during analyses to respect potential desires to refrain from reporting gender.

We defined enrollment in this study as the return of DNA samples and signed consent forms. T0 characteristics of interest included if enrollees were adopted, had a personal diagnosis of cancer, a diagnosis of cancer in the family, had ever experienced a heart attack or had a family member experience a heart attack. Family in these questions was defined as a close biological relative, such as a mother, father, son, daughter, aunt or uncle. T1 characteristics of interest included sex assigned at birth, sexual orientation, education, income, any past genetic testing, or any genetic testing in the family indicating an increased disease risk. Similar to the T0 survey, the T1 survey also asked about personal and family history of cancer or heart attack and proceeded to go into more detail to learn exactly which family members had experienced these conditions. However, because T1 survey completion was lower compared to T0, T0 survey data were used to report personal and familial cancer and heart attack status.

We calculated the number of study invitees and enrollees and descriptive statistics for study invitees and enrollees using EHR demographic data. Demographics of enrollees were further described using data from the T0 and T1 surveys. We also descriptively assessed overall drop out in this screening study by looking at the number of people who proceeded through different steps: (1) sent study invites, (2) accessed study FAQ, (3) requested a DNA collection kit, (4) sent a DNA collection kit and consent forms, (5) returned DNA sample and signed consent forms, (6) sent results. For the sent results step, we described the number of people with available screening results. We additionally reported the number of people with positive results that the study genetic counselor was able to reach to discuss results. We also analyzed enrollee activity on the study results online portal to determine the number of enrollees with uninformative results who accessed their result letter. Study drop out was also descriptively assessed by race and ethnicity.

We used two logistic regression models to examine the association between race and ethnicity and enrollment in population genetic screening. Model A included age (linear), gender (Male, Female, Other), and race/ethnicity (ascertained from the EHR: Hispanic (of any race), Asian [reference], African American, Multiracial/Other, Native American, White (latter five categories were all Non-Hispanic) as covariates. Model B additionally included a gender by race and ethnicity interaction term as differences in engagement in genetic services and research has been seen based on gender and race and ethnicity.^4-6^ We investigated significant interaction effects by calculating the probability of enrollment for individuals in groups relevant to the interaction using the mean age of invitees. We also conducted an exploratory analysis to assess the relationship of sexual orientation (a binary variable indicating if an individual was in the group of 1,000 LGBTQ+ invitees as coded in the EHR) with study enrollment by adding sexual orientation and a sexual orientation by race and ethnicity interaction term as additional covariates to the original Model A. Individuals who had selected “Prefer not to answer” for questions regarding gender in the T1 survey were excluded from these analyses.

*Screening yield*

We described the total number of enrollees with screening results, including the number of positive and uninformative screening results and the pathogenic and likely pathogenic variants detected. Using T1 survey data, we calculated the number of people with positive results finding out about their genetic variant for the first time through screening. Comparisons of self-report of testing between of groups receiving positive and uninformative results were performed using Fishers exact test.

*Assessment of personal and family history*

We separated enrollees into several categories according to self-reported personal and family history: those already aware of their variant, those who reported a first-degree relative with a variant, those with a personal diagnosis of disease, those with a first degree relative diagnosed with disease, and those with no reported family history of disease. The T0 pre-enrollment survey only asked a few high-level questions about personal and family history of cancer and heart disease. The T1 survey asked more detailed questions about personal and family history. For family history assessment, data from the T1 survey were used when possible.

*Assessment of guidelines*

We used self-reported personal and family history, including history of prior genetic testing to determine if the enrollees were likely to have qualified for genetic testing under National Comprehensive Cancer Network (NCCN) guidelines for testing for any of the cancer-risk genes on the panel or if they would have qualified for familial hypercholesterolemia testing under American College of Cardiology guidelines.^7,8^ Because personal and family history were self-reported and the study did not have access to complete medical records, it was only possible to evaluate a few of the guideline criteria. Most criteria for familial hypercholesterolemia diagnosis rely on lipid measures, which were not available to the study.

#### 3. **Study information and frequently asked questions provided by the population genetic screening study at UWM**

**Study Information**

**Q. What do I have to do to join the study?**

**A.** First, you have no obligation to join this or any research study. There are no negative consequences if you do not want to join. For any questions you can call Dr. Brian Shirts and the study team at (206)685-1176 or.

We must collect your DNA for testing. In a week or two, a saliva collection kit with instructions and consent forms will be sent to you via mail. Please read the consent forms carefully. Return the saliva kit and signed consent forms in the return, postage paid, envelope provided. We get less than a teaspoon of saliva samples. **We cannot receive the saliva sample or test your DNA without the signed consent forms.**

To learn more about this research study, please read the following information. You will be asked a few questions in the next few screens.

**Q. What information do you need?**

**A.** We need to know your address and phone number to send you a saliva DNA collection kit and consent form.

Once we receive your sample and consent form in the mail, we will ask you to fill out more questions to tell us about your feelings around genetic testing. This survey will take no more than 20 to 30 minutes to complete. You can skip any questions you would rather not answer.

**Frequently Asked Questions**

**Q. How long will the DNA results take?**

**A.** DNA research testing results take a while. It is our goal to get results to all participants within 6 months or sooner. After receiving your results, your participation is complete. However, you are free to contact the Researchers at any time.

**Q. What kind of results will I get?**

**A.** Most people will find that they do not have increased risk for inherited disease that can be identified by this screening test. Those people will get an e-mail allowing them to look up their results on a secure study website or can opt to receive a mailed letter with their results. While these results are reassuring, they do not mean that there is no disease risk; it means that there is no increased risk that can be detected by the genetic screening test used in this study.

About one or two percent of people will get a “higher-risk” result. These people will receive a phone call or e-mail from a study Genetic Counselor or the Investigator to discuss the results. This test just measures risk (or chance) of developing a disease. Not everyone with a “higher-risk” result is certain to develop the disease. Because we conduct a screening test that will not go in your medical records, the results should be confirmed with follow-up clinical testing through your doctor. Printable "higher-risk” result reports will be available to you on the secure website so that you can share them with your doctor.

**Q. What about other genetic studies?**

**A.** Everyone that participates in this study can also join the University of Washington – Brotman Baty Precision Medicine Institute (BBI) Biorepository and Registry Study. A repository collects and stores samples and data to be used for future precision medicine research. A registry is a list of names of people who don’t mind being contacted occasionally to hear of new research. If you are interested in joining the BBI Repository/Registry study you can indicate this on a separate paper consent form that you will receive with the saliva DNA collection kit.

People who have a “higher-risk” result will also be eligible to join a study to learn more about their genetic risk, and also involve family members who may share their genetic risk.

**Q. Will my information be kept confidential and safe?**

**A.** All information you provide us will be confidential and we will make every effort to keep your information and samples safe.

**Q. How long will my samples and information be saved?**

**A.** Your samples may be saved indefinitely.

**Q. What if I change my mind?**

**A.** You can contact us at any time to remove your information/sample from the study or tell us to not return results to you.

**Q. What are the risks for me joining this study?**

**A.** It may be unpleasant to collect a sufficient amount of saliva (spit) for the DNA test. Learning about your genetics can be emotionally stressful. Some people cannot predict how learning genetic information may make them feel until after they have received the results. Talking about private matters and personal feelings may make you feel uncomfortable or embarrassed. Another possible risk is loss of confidentiality or private information should a data breach occur. We will do our best to help you understand your genetic results and to keep your private information secure.

Although The Genetic Information Nondiscrimination Act of 2008 federal law (GINA) has been passed to prevent discrimination based on genetic information, it is possible that taking part in this study might make it harder to gain and/or keep employment or insurance. If you have any questions about your rights as a research participant, you can contact the University of Washington Human Subjects Division at (206)543-0098 or call collect at (206)221-5940.

**Q. What are the benefits for me joining this study?**

**A.** We cannot predict if you will receive personal benefit from participating in this research. Our hope is that you will gain information on your genetic risks for a screening list of inherited diseases. Your participation may benefit society by helping to advance genetic science. We are hopeful that future generations may benefit from the scientific and medical knowledge we gain, and that better methods will be put in place to start using genetic information to prevent disease.

**Q. Will I get paid for this study?**

**A.** No.

**Q. Does this study cost me anything?**

**A.** No. The genetic screening test is free. Any follow-up medical care will not be covered by the study.

**Q. Who is funding this research**

**A.** This is funded by the Brotman Baty Institute for Precision Medicine at the University of Washington.
